# Pharmacoepidemiology simulation study practices: A methodological review

**DOI:** 10.1101/2025.04.30.25326427

**Authors:** Ryan Muddiman, Florencia Ines Aiello Battan, John Tazare, Anna Schultze, Fiona Boland, Teresa Perez, Li Wei, Mary E. Walsh, Frank Moriarty

**Affiliations:** School of Pharmacy and Biomolecular Sciences, RCSI University of Medicine and Health Sciences Ireland; Facultad de Estudios Estadísticos, Universidad Complutense de Madrid, Spain; Department of Medical Statistics, London School of Hygiene & Tropical Medicine, London, UK; Department of Non-Communicable Disease Epidemiology, London School of Hygiene & Tropical Medicine, London, UK; Data Science Centre, School of Population Health, RCSI University of Medicine and Health Sciences Ireland; School of Pharmacy, University College London, London, UK

## Abstract

**Purpose:** Simulation studies are used in pharmacoepidemiology for evaluating inferential methods in a controlled setting, whereby a known data-generating mechanism allows evaluation of the performance of different approaches and assumptions. This study aimed to review simulation studies performed in pharmacoepidemiology.

**Methods:** We conducted a review of all papers published in the journal of Pharmacoepidemiology and Drug Safety (PDS) over the period 2017 to 2024. We extracted data on study characteristics and key simulation choices such as the type of data generating mechanism used, inferential methods tested and simulation size.

**Results:** Among 42 simulation studies included, 34 (81%) were informing comparative effectiveness/safety studies. 22 studies (52%) used simulation in the context of a clinical condition, and 36 (86%) used Monte-Carlo simulation. Inputs not derived from empirical data alone (n=22, 52%) or in combination with real-world data sources (n=19, 45%) were most often used for data generation. The complexity of simulations was often relatively low: although 31 studies (74%) generated data based on other covariates, time-dependent covariates (n=3) and effects (n=4) were rarely implemented. Bias was the most often used performance measure (n=26, 62%), although notably 18 studies (43%) did not report uncertainty in the method.

**Conclusion:** Simulations contributed a relatively small number of articles (3.2 % of 1320) to PDS over 2017 to 2024. Greater focus on evaluating methods and inferential approaches, using simulation studies that are appropriately complex given clinical realities may be beneficial to the pharmacoepidemiology field.

## Introduction

Simulation studies are empirical experiments that typically generate data from pseudo-random sampling, apply quantitative estimation, and evaluate the estimation method.(1) They are particularly useful when data complexity makes theoretical evaluation of methods impossible(2), such as estimating the power of a test given the data structure alone. This is particularly true in observational pharmacoepidemiology datasets where complex and dynamic processes are involved.(3) Simulations can evaluate inferential methods in a controlled setting, where aspects of the underlying data generating mechanism are controlled (e.g., surrounding treatment effect and confounding structure). In pharmacoepidemiological research, this can be applied to the estimands of interest, most often relative drug efficacy/safety or the prevalence of clinical outcomes.

Various approaches can be used in the DGM. Where real-world data (RWD) is available, the DGM can involve plasmode simulation,(4) which samples covariates from a real dataset and simulates outcomes, enhancing external validity. Monte-Carlo simulations(5,6) instead involve repeating simulations while varying input parameters to provide generalised results across scenarios. The approaches for simulation studies commonly used by the pharmacoepidemiology community have not been explored to date. Understanding current trends in the field could support our understanding of current best practice and highlight potential future areas for improvement. Therefore, this study aimed to review simulation studies and approaches used in pharmacoepidemiology.

## Methods

The protocol for this review was pre-registered(7) and the details are summarised below.

### Search

Our study adopted similar approaches to a previous review of code-sharing practices in pharmacoepidemiology and focused on papers published in the journal of Pharmacoepidemiology & Drug Safety (PDS)(8). We included simulation studies identified in this previous review (2017-2022) and augmented them with our own comparable search of the literature from January 2023 to December 2024. Thus, our study covers the period 2017-2024. PubMed was searched using the easyPubMed package(9) in R(10) (see the supplementary material for the search string). Our finalised search was conducted on 17/01/2025.

### Eligibility

Articles included were original research articles or brief reports where computer-simulated data was used to evaluate an inferential method. Commentaries and review articles were excluded. Studies that did not involve computer simulations (to avoid including pedagogical healthcare simulations) or did not measure the performance of an inferential method were also excluded. Reasons for full-text exclusion are given in the supplemental information.

### Screening

Covidence was used for screening of titles/abstracts of simulation studies from the previous review, and records from the new search, by two independent reviewers (RM, FAB), with conflicts resolved by a third reviewer (FM). Full-texts were reviewed by the same two independent reviewers, with conflicts and excluded records checked and resolved by the same third reviewer.

### Data extraction

All data extraction was performed by one reviewer and a second reviewer also extracted 20% of articles to assess consistency. For all eligible articles basic metadata was extracted such as the PMID and year using the easyPubMed package. The remaining data was extracted manually using an MS Excel spreadsheet with data validation based on the protocol(7). The type of data extracted was classified using the Aims, Data-generating mechanism, Estimand, Method, Performance measures (ADEMP) framework by Morris *et al*.(1). Where data items were not reported in the article, simulation code (if provided) was inspected to identify the missing item.

### Data analysis

We used R (version 4.4.1) to analyse extracted data, focusing on descriptive statistics. The code and data are on Zenodo.(11)

## Results

Forty-two studies were included (see supplementary material for the PRISMA(12) flow chart). Table 1 summarises the characteristics of included articles. Two were classified as brief reports and the rest were original articles as defined by PDS.

**Table 1.** Summary of characteristics of included simulation studies (n=42)

| Variable |  | n | % |
| --- | --- | --- | --- |
| Study type | Comparative effectiveness/safety | 34 | 81% |
|  | Signal detection | 4 | 10% |
|  | Exposure measurement | 2 | 5% |
|  | Covariate balance | 1 | 2% |
|  | Outcome rate estimation | 1 | 2% |
| Aspect of study simulation informed | Analysis | 29 | 69% |
|  | Design | 9 | 21% |
|  | Design and analysis | 4 | 10% |
| Type of intervention examined | Not applicable | 19 | 45% |
|  | Drug | 12 | 29% |
|  | Vaccine | 4 | 10% |
|  | No specific intervention | 4 | 10% |
|  | Multiple | 2 | 5% |
|  | Surgery | 1 | 2% |
| Drug class or vaccine | Not applicable | 26 | 62% |
|  | Other | 6 | 14% |
|  | Statin | 3 | 7% |
|  | Vaccine | 3 | 7% |
|  | Anti-cancer | 2 | 5% |
|  | Anticoagulant | 2 | 5% |
| Clinical condition examined<sup>1</sup> | No/Not applicable | 20 | 48% |
|  | Diseases of the circulatory system | 7 | 17% |
|  | Neoplasms | 5 | 12% |
|  | Endocrine, nutritional and metabolic diseases | 3 | 7% |
|  | Diseases of the respiratory system | 1 | 2% |
|  | Codes for special purposes | 1 | 2% |
|  | Diseases of the nervous system | 1 | 2% |
|  | Pregnancy, childbirth and the puerperium | 1 | 2% |
|  | Diseases of the digestive system | 1 | 2% |
|  | Diseases of the musculoskeletal system and connective tissue | 1 | 2% |
|  | Factors influencing health status and contact with health services | 1 | 2% |
| Year | 2017 | 4 | 10% |
|  | 2018 | 3 | 7% |
|  | 2019 | 10 | 24% |
|  | 2020 | 6 | 14% |
|  | 2021 | 3 | 7% |
|  | 2022 | 6 | 14% |
|  | 2023 | 5 | 12% |
|  | 2024 | 5 | 12% |
| Simulation type | Monte Carlo | 36 | 86% |
|  | Plasmode | 5 | 12% |
|  | Bootstrap<sup>2</sup> | 1 | 2% |
| Data generation mechanism | Random sampling | 36 | 86% |
|  | Stochastic process | 6 | 14% |
| Time parameterisation | None | 23 | 55% |
|  | Continuous | 15 | 36% |
|  | Discrete | 3 | 7% |
|  | Not reported | 1 | 2% |
| Source for data generation | Synthetic data | 22 | 52% |
|  | Both | 19 | 45% |
|  | Real world data | 1 | 2% |
| Factor variables<sup>3</sup> | Covariate magnitude | 24 | 57% |
|  | Effect magnitude | 18 | 43% |
|  | Data Generating Mechanism structure | 12 | 29% |
|  | Not applicable | 6 | 14% |
| Quantity of interest | Cumulative risk | 21 | 50% |
|  | Instantaneous risk | 12 | 29% |
|  | Other | 9 | 21% |
| Primary inferential method categorised | Estimation | 17 | 40% |
|  | Maximum likelihood | 9 | 21% |
|  | Partial likelihood | 11 | 26% |
|  | Other | 4 | 10% |
|  | NA | 1 | 2% |
| Performance measure for assessing inferential method | Bias | 26 | 62% |
|  | Other | 10 | 24% |
|  | Power | 3 | 7% |
|  | None | 3 | 7% |
| Uncertainty analysis | Summary dispersion | 19 | 45% |
|  | None | 18 | 43% |
|  | Coverage<sup>4</sup> | 4 | 10% |
|  | Other | 1 | 2% |
| Programming language | R | 20 | 48% |
|  | Unknown | 9 | 21% |
|  | Stata | 5 | 12% |
|  | R, & SAS | 4 | 10% |
|  | SAS | 3 | 7% |
|  | MATLAB | 1 | 2% |
| Published code | No | 24 | 57% |
|  | Yes | 18 | 43% |
| Component of code shared | Not applicable | 24 | 57% |
|  | Data generating mechanism and analysis | 11 | 26% |
|  | Analysis only | 5 | 12% |
|  | Data Generating Mechanism only | 2 | 5% |
| Location of shared code | NA | 22 | 52% |
|  | Supplemental | 13 | 31% |
|  | GitHub | 2 | 5% |
|  | Code package | 2 | 5% |
|  | Package | 1 | 2% |
|  | GitHub, supplemental | 1 | 2% |
|  | GitLab | 1 | 2% |

<sup>1</sup> The condition was classified according to the ICD-10 chapter.
<sup>2</sup> This study utilised full bootstrapping to obtain the data, whereas plasmodes contain covariate sampling with simulated outcomes.
<sup>3</sup> Factors are variables that are systematically varied through repeated simulations. This does not sum to the number of studies since more than one variable may be factored.
<sup>4</sup> Coverage is a performance measure relating to correct interval estimation and as such is classed here within uncertainty analysis.

The results of binary parameters are shown in Figure 1 (a). Twenty-three studies (55%) focused on a specific clinical question with the more common condition studied being diseases of the circulatory system (17%). Most simulations aimed to inform comparative effectiveness/safety studies (81%) and specifically their analysis (69%). Where medical interventions were named, drugs were the most common type (29%).

**Figure 1.**
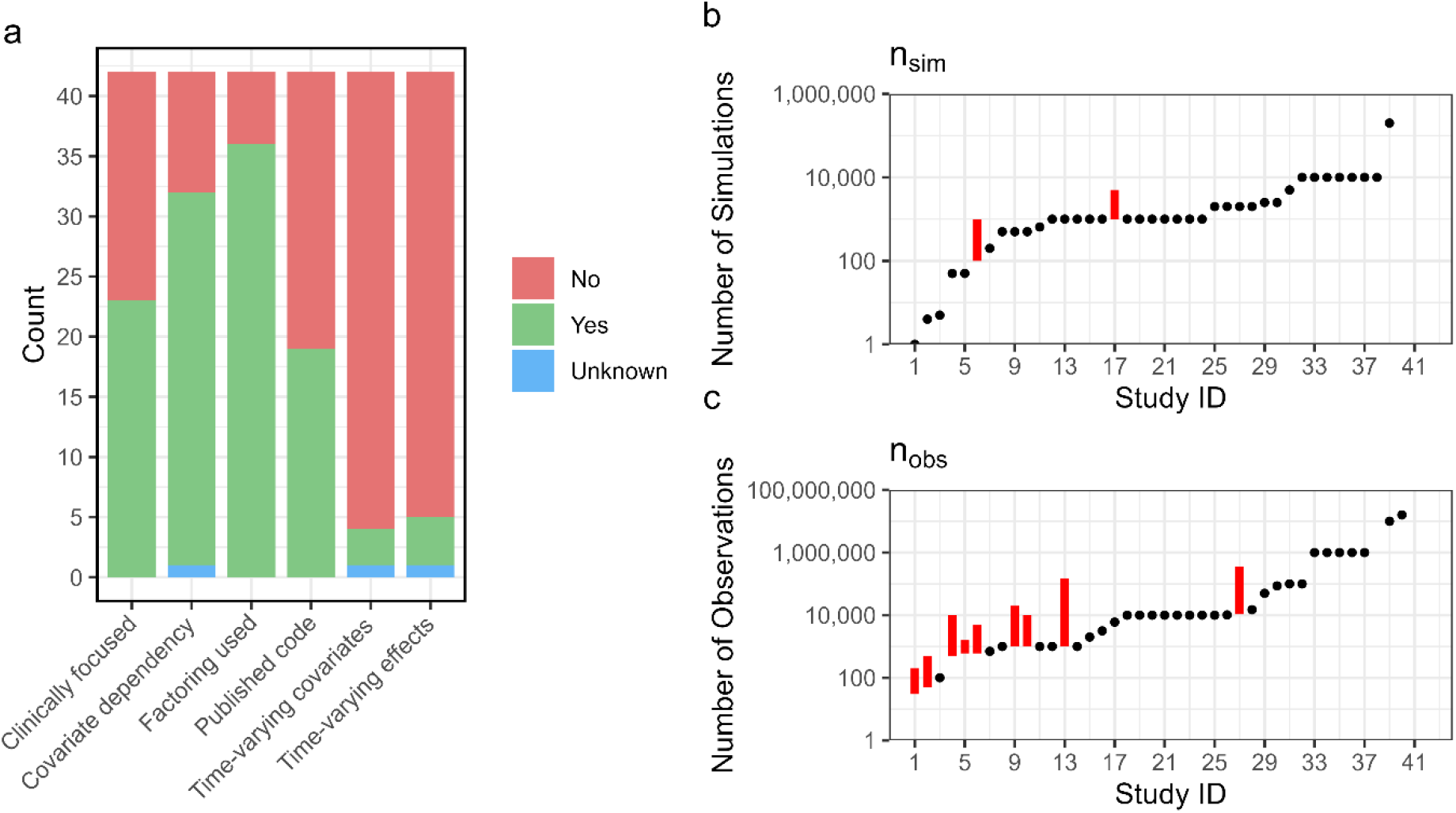
(a) Bar chart of binary study characteristics, (b) Distribution of values used across studies for the number of simulations (b) and number of observations (c). Study ID is not equal between (b) and (c). (Red bar is a range).

Simulations consisted of Monte-Carlo type (88%), plasmodes (12%) and bootstrap (2%). The DGM was characterised as random sampling in thirty-six studies (86%) and by a stochastic process in the remaining studies (random sampling includes time-independent outcome generation and DGMs were classified as stochastic where they generated a random variable indexed by a separate time variable). Fifteen studies (36%) had time defined continuously and three (7%) used a discrete time scale. The source for the DGM parameters was most frequently inputs not based on empirical data (52%) with other studies using only RWD or a mix of both.

Thirty-one studies (74%) had generated data that had exposures or outcomes dependent on other covariates and thirty-six studies (86%) performed factoring of one or more variables in the simulation. The most commonly factored parameters were the magnitude of covariates (57%). Three studies (7%) included time-varying covariates and four (10%) included time-varying effects in simulations, although time was not a relevant aspect of some studies. We show the distribution (over all studies) of the minimum and maximum of the number of simulations (n_sim_) and number of observations (n_obs_) per simulation in Figure 1 (b) and (c) respectively. The number of simulations was mostly defined as a single value (88%), while the number of observations was varied more frequently (24%).

The most common target in the simulations were risk measures (50%) such as an odds ratio or relative risk, other measures such as hazard ratios were less common (29%). Nonparametric estimation was the most common inferential method used (40%). Other methods used maximum and partial likelihoods. Twenty-six studies used bias as the primary measure of performance (62%), and the uncertainty was mostly reported using dispersion measures over the simulations such as confidence intervals and standard deviations (45%). The most common programming language was solely R (48%); however, 9 studies (21%) did not indicate the software used. Code was shared in less than half of studies (43%). The shared code was primarily used for both the data generation mechanism and analysis (26%) but some code only performed one of these functions. The shared code was in the supplemental information of 13 studies (31%).

## Discussion

Simulation studies represented a very small number of articles in PDS over 2017-2024 (3.2%). Studies mostly used pseudo-random sampling for outcome generation within a Monte-Carlo simulation framework incorporating synthetic covariates. The Monte-Carlo simulation generates multiple pseudo-observations which are then analysed using the inferential method of choice. Some studies lacked reporting of key simulation details such as the software used and choice of time definition in the DGM. Despite minimal legal/regulatory impediments, code was often not shared, limiting replication of studies using different methods.

Over half of studies focused on a clinical scenario such as a specific drug class, allowing potential DGM inputs to be narrowed to those expected in reality. In such cases, relative risks or other causal or associational measures may be based on real data or expert judgement. When the simulation is not based on a clinical scenario, the choice of input distributions may be less clear, and may result in selections that are unrealistic in practice. Comparing the simulated dataset to RWD using summary statistics is a potential solution.

Plasmodes were uncommon, potentially due to the lack of access to RWD which requires ethical approval and also lack of guidance for implementation. Alternatively, published RWD effect estimates can be used in a Monte-Carlo simulation with synthetic covariates, although typically only the summary statistics of covariate distributions are published. Observation numbers are restricted to the available data in a plasmode or when using RWD. In Monte-Carlo studies, the number of observations is determined by the researchers and thus varied substantially, with a median of 10,000 for included studies. Conversely, the number of simulations is not bounded by any external data. While a higher value of n_sim_ will reduce the sampling error of Monte-Carlo estimates, there was typically no justification given for the actual choice of n_sim_, and in practice this often relates to available computing resources. However, uncertainty in the estimate was often not reported (n=18, 43%), meaning the sampling variation due to the finite number of simulations cannot be ascertained.

Time-constant effects and covariates were simulated in four and three studies respectively. The choice of time-constant parameters means that the simulated data represented a static risk/relative risk of outcomes over time (i.e., meeting the proportional hazards assumption), which may not represent reality in some scenarios.(13) While some studies did evaluate multiple statistical methods and reported different performance measures, it was previously observed that the relative performance of different statistical methods depends on the DGM structure and the type of distributions used(14) and at a minimum, simulations should incorporate both time-constant and time-varying effects, and dependence on past events. Thus, DGMs were often not adequately varied or suitably complex for the conclusions of many studies to be applicable to observational datasets.

While this review was limited to a single journal, it suggests low numbers of simulation studies published within the pharmacoepidemiology community. Greater focus on evaluating methods and inferential approaches, using simulation studies that are appropriately complex given clinical realities, may be beneficial to the field.

## Data Availability

All data produced are available online at https://zenodo.org/records/15309637

## Notes

### Competing Interest Statement

AS is employed by LSHTM on a fellowship funded by GSK.

### Clinical Protocols

https://doi.org/10.17605/OSF.IO/QZSAR

### Funding Statement

This work was supported by the Wellcome Trust (DIAMOND programme Career Development Award, grant number 227348/Z/23/Z). TP was funded by PID2022-137050NB-I00 of the Spanish Ministry of Science and Innovation.

