## Supplementary information for "Pharmacoepidemiology simulation study practices: A methodological review"

*S 1 Flow chart for search*

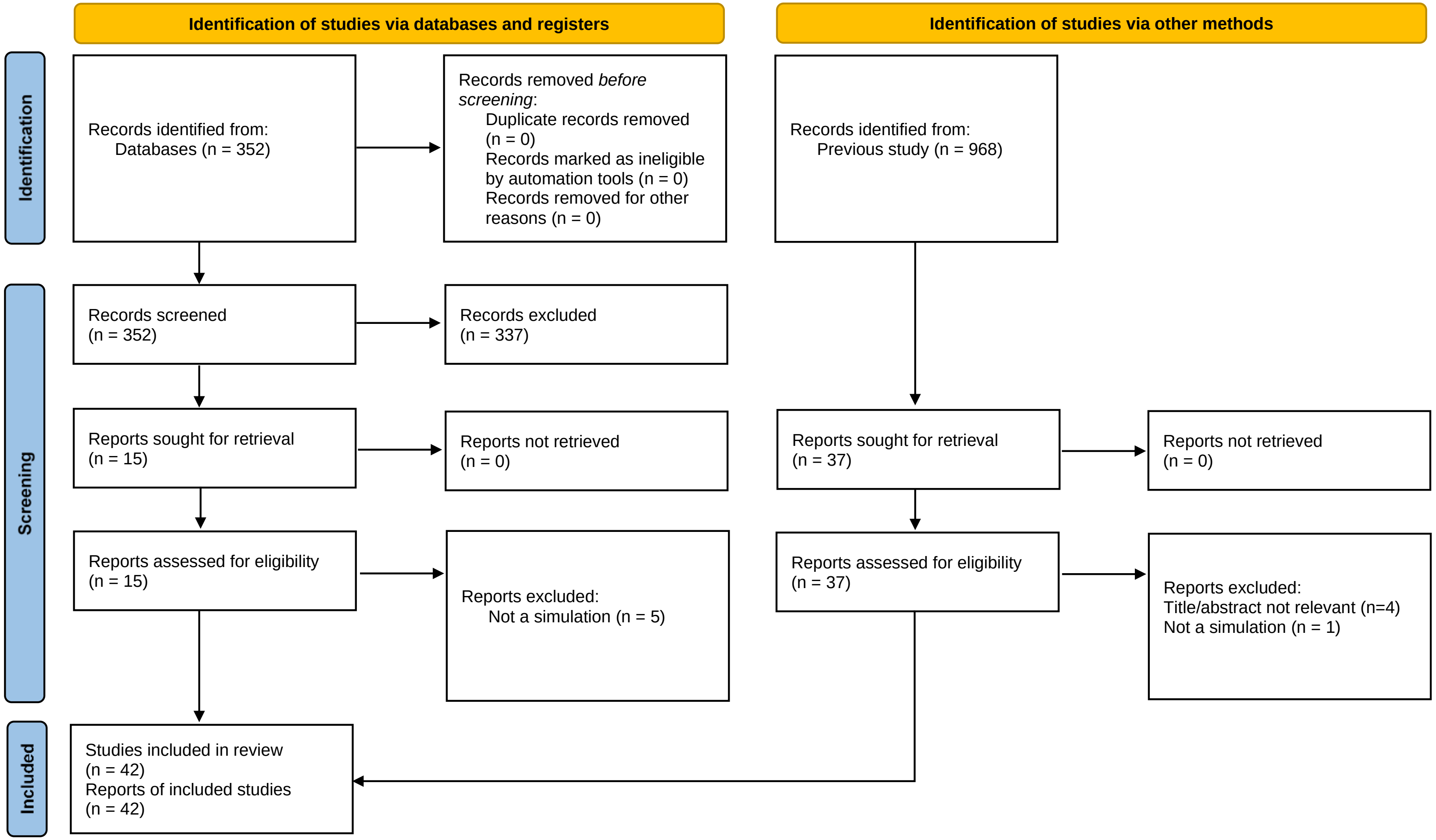

*S 2 Table of excluded articles from full-text stage*

| PMID | Title | Reason for exclusion |
|---|---|---|
| 37609668 | Combining Super Learner with high-dimensional propensity score to improve confounding adjustment: A real-world application in chronic lymphocytic leukemia | No inferential method was tested, true effect was not known |
| 37528702 | Importance of accounting for timing of time-varying exposures in association studies: Hydrochlorothiazide and non-melanoma skin cancer | No inferential method was tested, true effect was not known |
| 39013838 | Optimization-Based Stable Balancing Weights Versus Propensity Score Weighting for Samples With High Covariate Imbalance. | No inferential method was tested, true effect was not known |
| 37850535 | How to account for early overly small risk sets in the analysis of pregnancy outcome data?-Comparison of different methods for stabilizing the Aalen-Johansen estimator. | No inferential method was tested, true effect was not known |
| 38453354 | An open-source implementation of tree-based scan statistics | No inferential method was tested, true effect was not known |
| 32394495 | The impact of statin discontinuation and restarting rates on the optimal time to initiate statins and on the number of cardiovascular events prevented | No inferential method was tested, true effect was not known |

### Search string

The search in PubMed consisted of the following string

' "Pharmacoepidemiology and drug safety"[Journal] AND (("2022/12/31"[Date - Publication] : "2024/12/31"[Date - Publication]))'.

### Data extracted

Data relating to simulation aims were clinical relevance (yes/no), clinical condition examined (classified using ICD-10 chapter), the type of intervention studied, and the specific drug class studied. The DGM aspects that were extracted included the simulation type (Monte-Carlo/Plasmode/other), DGM (stochastic process/random sampling), time parameterisation (continuous/discrete/none), whether outcome data depended on covariates (yes/no), whether there were; time-varying effects (yes/no), time-varying covariates (yes/no), time-varying effects (yes/no), the source for the data generation (RWD/synthetic/both), whether or not factoring was used (yes/no) and the variables that were factored. We also extracted the number of simulations used and number of observations per simulation. The data relating to the simulation estimand was the primary estimand of the simulation. We also extracted the performance measure used and how its uncertainty was quantified. Finally, we extracted data on ancillary information such as the programming language used, whether the code was shared publicly and what the shared code was used for (DGM/analysis/both).

Three extracted items were added after registration of the protocol. This was the type of research study that the simulation was aimed at informing (study aim), the aspect of this study that would benefit from simulation (simulation focus) and the location of shared code. For the study aim, we categorised the clinical focus of the simulation only. Some studies presented an empirical application of a method on RWD, but typically independent of the simulation.

*S 3 Data extraction table*

| **Variable** | | **Notes** |
|---|---|---|
| Study type | Comparative effectiveness/safety | e.g. the relative risk of a drug vs. a placebo for adverse events |
| | Signal detection | e.g. the prevalence of drug-drug interactions/adverse events |
| | Exposure measurement | e.g. classifying the value of exposure under uncertainty |
| | Covariate balance | e.g. propensity score matching |
| | Outcome rate estimation | e.g. Absolute risk |

| Variable | | Notes |
|---|---|---|
| Aspect of study simulation informed | Analysis | The simulation informed the type of analysis e.g., model selection |
| | Design | The simulation informed the design of a study e.g., classification of exposure/optimal sample size |
| | Design and analysis | The simulation informed the design and analysis of a study |
| Type of intervention examined | Not applicable | |
| | Drug | |
| | Vaccine | |
| | No specific intervention | |
| | Multiple | |
| | Surgery | |
| Drug class or vaccine | Not applicable | |
| | Other | |
| | Statin | |
| | Vaccine | |
| | Anti-cancer | |
| | Anticoagulant | |
| Clinical condition examined | No/Not applicable | Classified according to ICD-10 chapter |
| | Diseases of the circulatory system | |
| | Neoplasms | |
| | Endocrine, nutritional and metabolic diseases | |
| | Diseases of the respiratory system | |
| | Codes for special purposes | |
| | Diseases of the nervous system | |
| | Pregnancy, childbirth and the puerperium | |
| | Diseases of the digestive system | |
| | Diseases of the musculoskeletal system and connective tissue | |
| | Factors influencing health status and contact with health services | |
| Year | 2017 | |
| | 2018 | |
| | 2019 | |
| | 2020 | |
| | 2021 | |
| | 2022 | |
| | 2023 | |
| | 2024 | |
| Simulation type | Monte Carlo | Sampling from parametric models |
| | Plasmode | Sampling primarily from RWD (e.g. covariates) |

| Variable | | Notes |
|---|---|---|
| | Bootstrap [1] | Sampling with replacement from a fixed dataset |
| Data generation mechanism | Random sampling | e.g. Bernoulli trial for outcomes |
| | Stochastic process | DGMs were classified as stochastic processes where the outcome was generated as a random variable indexed by a separate time variable e.g. times of recurrent events |
| Time parameterisation | None | Time was not part of the simulation data structure |
| | Continuous | Time was specified continuously (e.g. using the inverse transform method) |
| | Discrete | Time was specified in discrete intervals (e.g. using a discrete Markov chain) |
| | Not reported | Study appears to involve time but the parameterisation was not reported |
| Source for data generation | Synthetic data | The study did not use RWD as an input or as a dataset for bootstrapping/sampling |
| | Both | The study used both RWD and user-specified data |
| | Real world data | The study used RWD as an input or in bootstrapping/sampling |
| Factor variables[2] | Covariate magnitude | e.g. Mean of parametric distribution representing a confounding variable |
| | Effect magnitude | e.g. True odds ratio of effect |
| | Data Generating Mechanism structure | e.g. number of confounders |
| | Not applicable | |
| Quantity of interest | Cumulative risk | |
| | Instantaneous risk | |
| | Other | |
| Primary inferential method categorised | Estimation | |
| | Maximum likelihood | |
| | Partial likelihood | |
| | Other | |
| | NA | |
| Performance measure for assessing inferential method | Bias | |
| | Other | |
| | Power | |
| | None | |
| Uncertainty analysis | Summary dispersion | |
| | None | |
| | Coverage[3] | |
| | Other | |
| Programming language | R | |
| | Unknown | |
| | Stata | |
| | R, & SAS | |
| | SAS | |

| **Variable** | | **Notes** |
|---|---|---|
| | MATLAB | |
| Published code | No | |
| | Yes | |
| Component of code shared | Not applicable | |
| | Data generating mechanism and analysis | |
| | Analysis only | |
| | Data Generating Mechanism only | |
| Location of shared code | NA | |
| | Supplemental | |
| | GitHub | |
| | Code package | |
| | Package | |
| | GitHub, supplemental | |
| | Gitlab | |
